# The same but different. Multidimensional assessment of depression in students of natural science and music

**DOI:** 10.1101/2023.01.10.23284333

**Authors:** Michaela Korte, Deniz Cerci, Roman Wehry, Renee Timmers, Victoria J. Williamson

**Affiliations:** The University of Sheffield, Department of Music, Sheffield, United Kingdom; Universitätsmedizin Rostock, Klinik für Forensische Psychiatrie, Zentrum für Nervenheilkunde, Rostock, Germany; Helios Klinikum Hildesheim, Akademisches Lehrkrankenhaus der Medizinischen Hochschule Hannover, Hildesheim, Germany; Independent Academic

## Abstract

Depression is one of the most common and debilitating health problems, however, its heterogeneity makes a diagnosis challenging. Thus far the restriction of depression variables explored within groups, the lack of comparability between groups, and the heterogeneity of depression as a concept limit a meaningful interpretation, especially in terms of predictability. Research established students in late adolescence to be particularly vulnerable, especially those with a natural science or musical study main subject. This study used a predictive design, observing the change in variables between groups as well as predicting which combinations of variables would likely determine depression prevalence. 102 under- and postgraduate students from various higher education institutions participated in an online survey. Students were allocated into three groups according to their main study subject and type of institution: natural science students, music college students and a mix of music and natural science students at university with comparable levels of musical training and professional musical identity. Natural science students showed significantly higher levels of anxiety prevalence and pain catastrophizing prevalence, while music college students showed significantly higher depression prevalence compared to the other groups. A hierarchical regression and a tree analysis found that depression for all groups was best predicted with a combination of variables: high anxiety prevalence and low burnout of students with academic staff. The use of a larger pool of depression variables and the comparison of at-risk groups provide insight into how these groups experience depression and thus allow initial steps towards personalized support structures.

## Introduction

Depression is one of the most common and debilitating mental health problems worldwide (1). The diagnosis and treatment of depression present specific challenges in different age groups, and one critical period is late adolescence (2,3). Moreover, students in higher education have been found to be significantly more affected by depression compared to peers pursuing a different educational route. Depression causes students additional distress during an already vulnerable time in their development (4). It is also associated with substantial impairment in academic performance and has the potential to cause lifelong problems (5,6).

Depression research in students has accumulated a wealth of information by focusing investigations on particular student groups (e.g. science students), but comparisons between student groups from different fields are scarce. While there is, for instance, solid evidence that medical students are significantly more affected by depression than the general population or their peers from different fields of study (7), little information has been gathered on students of music. Yet, the latter seem to be exceptionally predisposed to depression (8,9). Another key difference between the research in the two fields is that while studies with natural science students have based their evidence on a large pool of variables (e.g. anxiety, burnout), for music students, the focus has been mainly on performance anxiety and a historical deduction that depression is key for creativity. Evidence, however, points to the contrary yet, the belief in the necessity of mental suffering to heighten creativity remains entrenched (10). Research of depression in both medical students and music students claims that each groups’ specific stressors and environments create a uniquely vulnerable group, more affected by depression compared to others. The few existing comparison studies between medical and music students found no significant difference in terms of depression and anxiety (11). This suggest that both groups might have common denominators to depression that so far have not been investigated due to the specificity of the research.

Despite its ubiquitous reach, the heterogeneity of depression makes it difficult to diagnose and treat (12). With the progression of advanced testing, many diseases that had previously been clustered into one, have now been separated into more elucidated etiologies and can thus be viewed more objectively regarding their heterogeneity. This, however, is not the case for depression. Here, we can solely observe ‘a syndromic constellation of symptoms that hang together empirically, often for unknown reasons’ (13). This has been demonstrated by Østergaard et al.’s (14) mathematical demonstration of 1497 combinations of depression symptoms. This is not mere theoretical assumption but supported by research in the variability of depression trajectory and treatment variability (16). One way forward could be using different modeling techniques, using depression symptoms and patterns as predictors, thus testing in-connectivity or resilience to depression in (specific) groups. However, larger models and comparing different participant groups to elucidate patterns and improve predictability has not been widely adopted in the medical field, despite its demonstrated potential in cardiology and psychiatry (17,18).

This study will thus combine both approaches to identify commonalities and differences between groups of science students (including medicine) and music students. It will investigate depression through the lens of six depression variables (anxiety, depersonalization, coping strategies, professional identity, pain catastrophizing and burnout), firstly as individual variables and secondly as predictors using their cumulative weight. Depression and pain research has shown that different individuals can perceive pain to a similar degree and yet react to it differently. In order to be a valid variable/predictor in depression, pain needs to be processed in a dysfunctional way (19). Thus, we used pain catastrophizing as variable and pain perception as additional information. High anxiety has been individually linked to student groups in depression research (7,11). Correlations of depression with anxiety, depersonalization, and burnout symptoms suggest a comorbidity or, at least, common neurobiological denominators (20). Clinically observed phenomena, such as the activation of prefrontal attentional brain systems, present compelling evidence that links depersonalization and anxiety as co-morbid disorders (21). On the surface, it is easy to confuse burnout with depression as burnout is a syndrome that results from chronic workplace stress that has not been successfully managed. Psychological pressure on students can lead to emotional fatigue, poor personal performance, and depersonalization (20,22). High professional identity has been hypothesized as a potential depression variable for students following a vocational path such as music students. Professional identity has been found to increase depression, namely when a career change is necessary (e.g. after an injury), further isolating individuals by uprooting them from their social circle (23,24).

This study will compare three student groups: (1) science students (e.g. medicine) with no interest in music, (b) a mix of science students, who had initially focused on studying music and music students at university, and (c) music students at music college. It will be guided by three individual questions: (1) how does depression prevalence compare between groups? (2) how do depression predictors (variables) compare between groups? and (3) what role, if any, does professional identity as a musician play in depression? To answer these questions, the study will determine which known factors influence reported depression, how or if they differ between groups, and if the level of professional musical identity can be considered a predictor variable for depression. The outcomes of this study will provide timely evidence on how we may better identify students who are at risk of depression during their training.

## Materials and Methods

This quantitative study followed a predictive design, observing the change in variables between groups as well as predicting which combinations of variables would most likely determine a high depression prevalence. Using random sampling, data was obtained via an online survey. Participants had to be at least eighteen years of age and studying in higher education.

### Procedure

Participants were recruited online via the students’ servers. This study was carried out in accordance with the recommendations of the university and the protocol was approved by the Ethics Committee of the Department. All subjects gave written informed consent.

### Material

Participants were asked to share demographic details such as age, relationship status, etc (see table 1).

**Table 1.**
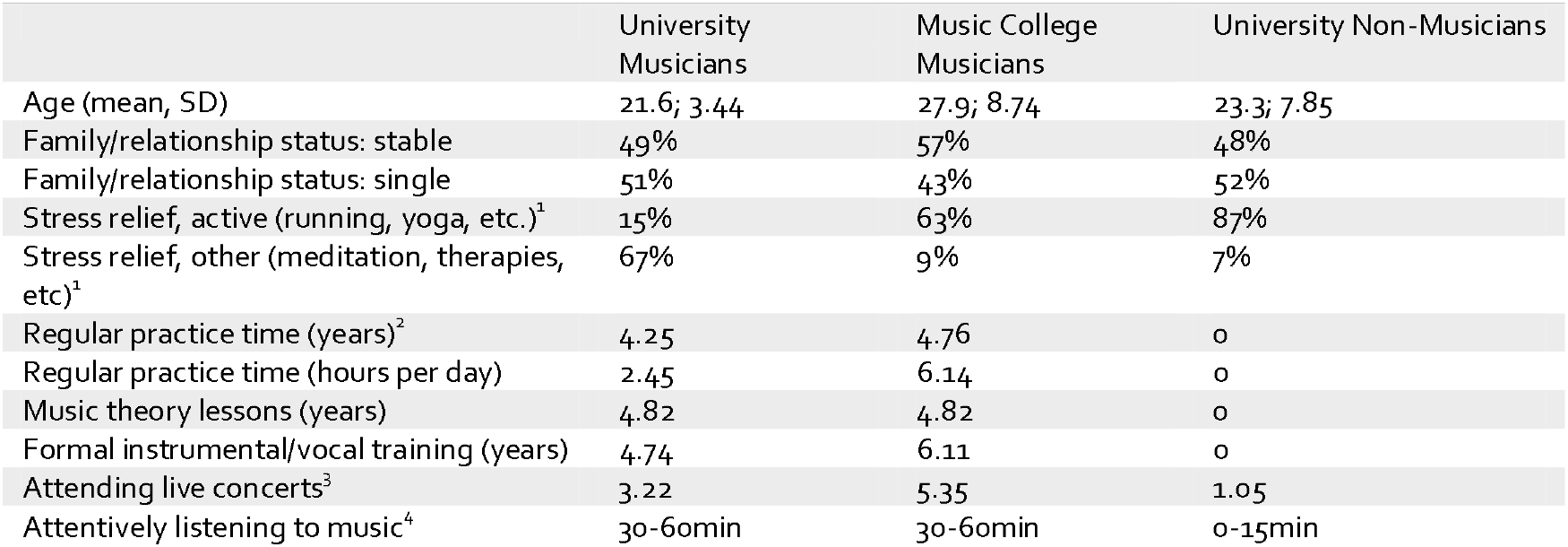
Demographics.

The **Hospital Anxiety and Depression Scale** (HADS) (25) is a self-reported questionnaire. It collects information on depression and anxiety symptoms using two separate scales (7 items per scale) based on a 4-point Likert response (Chronbach’s α□=.6 (26)). The HADS discriminates well between anxiety and depression (27). It is a good fit to the Rasch Model, stable across professions and less vulnerable to cultural bias (26).The cut-off for significant depression and anxiety was set at *≥* 9 (28).

The **Cambridge Depersonalisation Scale** (CD-9) (29) covers depersonalization through 9 questions. This scale has shown adequate internal consistency and temporal stability (α□= .92, retest reliability 10-14 days: *r*_tt_ = .86). Scores are added up and can reach 0 – 90, with 0 indicating no depersonalization. The cut-off point for significance was set at the level requested by the scale’s authors at ≥ 19 (short, transient) and ≥ 90 (unique condition).

The **Örebro Musculoskeletal Pain Screening** (30) measures how participants’ experience of pain affects their performance at work/higher education. In 21 questions it addresses pain beliefs and expectations. The higher the final score, the less likely the individual is to return to work while remaining disabled by pain. The authors specified that, using a six-month prediction, 71% of patients were correctly classified (sensitivity, 72%; specificity, 70%), with a high reliability (α = .97, *p* □ ≤.05)) and a high internal consistency (α = .87). Total score of 105 indicate a moderate risk, ≥ 130 a high risk of being disabled by pain.

The **Brief COPE** (31) distilled the 14 scales from the original questionnaire into three scales (28 questions). This allows for diverse testing of stress coping and correlation of findings. The three scales are: active functional coping (e.g. ‘I actively did something’), functional cognitive coping (e.g. ‘I tried to find something positive in what happened.’) and dysfunctional coping strategies (e.g. ‘I used alcohol/other substances to help me through this situation’). Internal consistency was found to be good for all subscales: emotion-focused, problem-focused, and dysfunctional subscales (α = 0.72, 0.84, 0.75)(32)

The **Copenhagen Burnout Inventory**, (CBI (33) consists of three main scales: personal burnout, work burnout, and client-related burnout. The authors’ attested all three scales to have very high internal reliability (α = .85 - 87). This study’s design was modelled on the study by Campos *et al*. (34) to reflect the dual ‘client’ burnout problem of students: the ‘client’ questions were doubled up, exchanging the word ‘client’ with ‘fellow student’ in one set, and ‘professor’ in the other. These scores reflect the level of exhaustion and fatigue perceived from this interpersonal relationship that derives from the students’ interaction with fellow students and/or academic staff (professor).

The **Athletic Identity Measurement Scale** (35) is a 10-item scale that assesses the strength and exclusivity of professional identity. The higher the score, the more a candidate identifies with being an athlete (10 – 70, mean of 40; internal consistency of *r* = .93; test-retest reliability of *r* = .89 (36)). We used Vitale’s (37) adaptation for musicians, changing the word athlete to musicians and called the questionnaire Musicians Identification Measurement Scale (MIMS).

The **Goldsmiths Musical Sophistication Index** (Gold-MSI) (38) is a self-reported test that assesses an individual’s propensity to engage with music. It is modelled on multidimensional construct of musical sophistication. With Chronbach’s α = .914 the scale is suitable instrument (38). We used two of the test’s subscales: active engagement and musical training (7 and 9 questions).

### Statistical analysis

Evaluation of the data was performed using RStudio 1.2.1335 (39) and G*Power (40). Power of *β* = .8 was considered as appropriate (41). Power calculations found the minimum for pairwise comparison with an expectation of non-linear distribution to be 96 participants in total (or 33 participants per group), and a minimum of 82 participants for a linear multiple regression for a fixed model. Missing values were imputed using the package mice (42), Party was used for the tree model (43). Chi square, Mann-Whitney U-test and ANOVA were used for (pairwise) comparisons. Regressions and a tree model were performed to analyse group differences and predictability of depression. Bonferroni corrections were applied to safeguard against multiple testing, and Spearman’s correlation for correlations.

## Results

102 under- and postgraduate students (75% United Kingdom, 16% other European Union countries, 9% United States of America; age mean = 23.6 years) from various institutions and with different primary study subjects (62% music, 38% medicine, psychology, biology) participated in this study. 67 students were from the University of Sheffield and 36 students from various music colleges. The students from music colleges remained together in one group, while the group of university students was made up of two sub-groups: 31 musicians and 36 non-musicians. The musicians’ group comprised university students who self-identified as musicians irrespective of their main study subject (music or science). The musicians’ group comprised university students who self-identified as musicians irrespective of their main study subject (music or natural science). Participants in this group showed equal levels of practice, lessons taken and engagement with music as the group of students from music college (see table 1).

Participants in the non-musician group (science) showed hardly any engagement with music (no instrumental or theory lessons). For ease of reference, the three different groups will from now on be referred to as university musicians, university non-musicians and music college musicians (see all tables below).

### Scale Outcomes

#### Depression and anxiety

There was a significant difference in depression prevalence between the music college musicians’ group and both the university musicians (*z* = -3.67, *p* = .0002), and the university non-musicians (*z* = 2.16, *p* = .003). Music college musicians showed significantly higher depression prevalence compared to university musicians and non-musicians (see table 2). The highest anxiety prevalence was found in university non-musicians. When compared to music college musicians, the difference was significant (*z* = -2.01, *p*= .04). There was no statistically significant difference between university non-musicians and university musicians (*p* = .2).

**TABLE 2.**
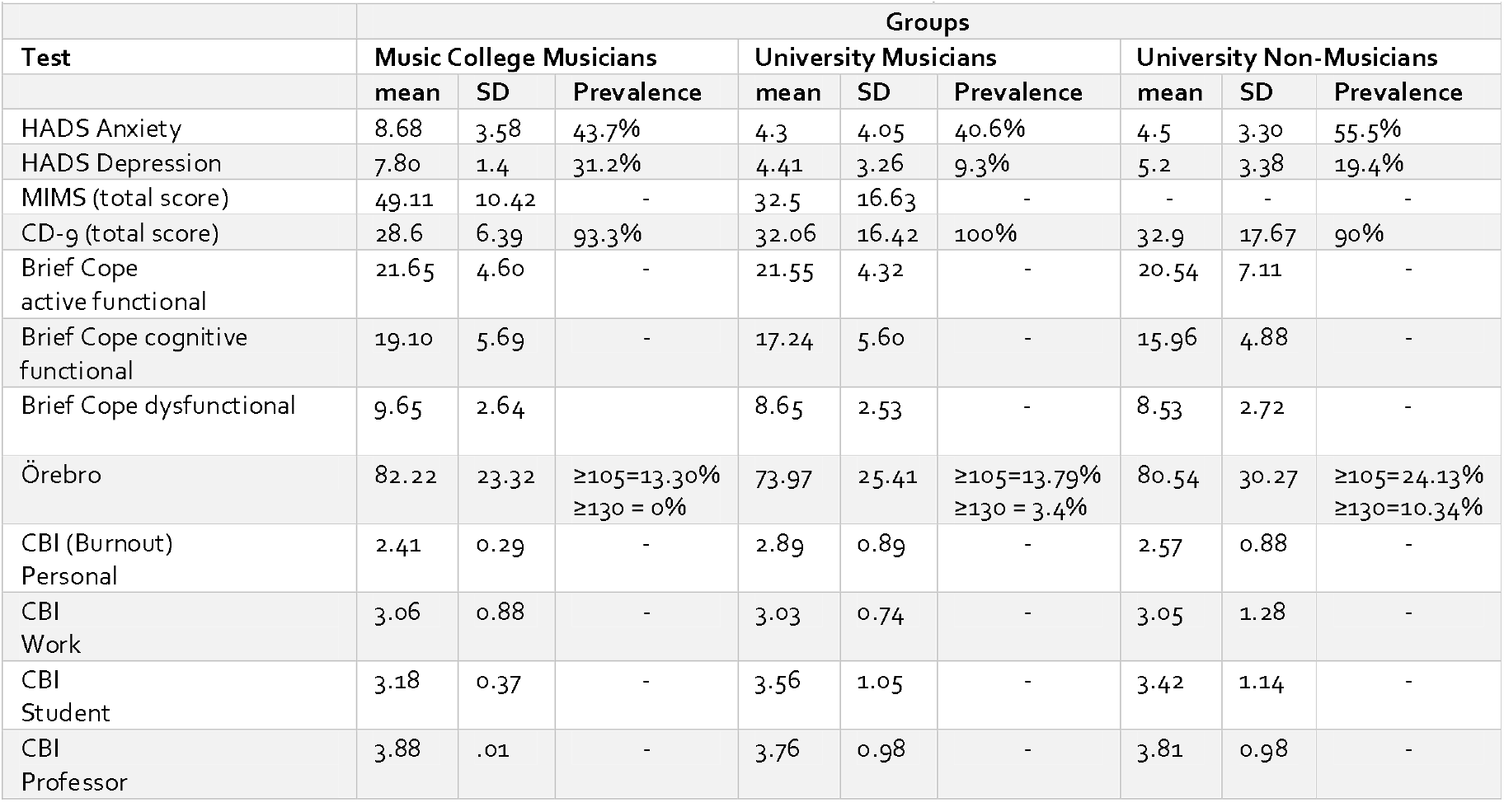
Results from all standardized tests (mean, standard deviation (SD) and prevalence)

#### Pain catastrophizing

University non-musicians showed a significantly higher prevalence of pain catastrophizing (≥130) compared to music college musicians (*z* = 1.94, *p* = .05). There was no statistically significant difference between university non-musicians and university musicians (*p* = .3).

#### Depersonalization

There was almost no difference in depersonalization prevalence between college musicians (93.3%), university musicians (100%) and university non-musicians (90%).

#### Coping

An ANOVA found no significant difference between groups for this variable (active functional cope: *p* = .6; cognitive functional cope: *p* = .2; dysfunctional cope: *p* = .7).

#### Burnout

There was no statistically significant difference between groups. The highest level of burnout was experienced based on interactions with teaching staff, followed by fellow students, personal burnout and then work burnout.

### GOLD-MSI and MIMS

Music college musicians invested more time into daily practice and formal lessons than university musicians, but did not accumulate more years of practice. Music college musicians spent more time listening attentively to music and attended more live concerts (audience) than university musicians. Non-musicians took no instrumental or theory lessons. They listened less to music and attended fewer concerts (see table 2). Moving on to the MIMS, a Mann-Whitney U-test determined a significant difference in the full score between music college musicians and university musicians, with a large effect size (*U* = 856.0, *p* = .001, rank-biserial correlation = .57). The subscales self-identity (*U* = 853.0, *p* < .004) and social identity (*U* = 588.0, *p* < .004) were significantly higher. Negative affectivity (*p* = .4) and exclusivity (*p* = .5) did not differ significantly.

### Modelling

#### Hierarchical linear multiple regression

The overall multiple linear regression model, using (HADS) anxiety, pain catastrophizing, professor burnout and the MIMS full score, was significant *F*(4,86) = 34.05, *p* <.001, *R*^2^ = .595. The multiple linear regression model, comprising all independent variables but MIMS was equally significant: *F*(3,87) = 33.33, *p* <.001, *R* = .535. All individual variables were significant: anxiety (*b* = .58, *t*(87) = 10.7, p < .001, *pr*^2^ = .568), pain catastrophising (*b* = .015, *t*(87) = 1.67, *p* < 001, *pr*^2^ = .031) and professor burnout (*b* = -.25, *t*(87) = -1.72, *p* < 001, *pr*^2^ = -.035). Anxiety emerged as the strongest depression predictor. With every .58 unit increase in anxiety the model predicted one unit increase in depression. Both pain catastrophizing and professor burnout predicted depression with approximately similar strength, but both were weaker predictors compared to anxiety. Burnout returned an inverse score. The question for this calculation was if adding the variable musical identity (MIMS) would significantly improve the model. The comparison of both models with an ANOVA showed that this was not the case: Δ*F*(1,86) = 2.50, *p* = .11. Musical identity was not significant within the model (*p* =.1). Moreover, the levels of predictability of the individual variables within the model changed with the addition. While anxiety and professor burnout increased in importance (anxiety: *b* = .58, *t*(86) = 10.8, *p* < .001, *pr*^2^ = .575; professor burnout: *b* = -.43, *t*(86) = - 2.34, *p* = .02, *pr*^2^ = -.06), pain catastrophizing became an insignificant predictor (*p* =.06). In summary, the hierarchical linear regression model does not support the hypothesis that musical identity is a valid predictor for depression.

#### Tree model

For the tree model, we calculated a decision tree to predict depression using all variables above (tab 1 and 2): The results can be seen in Figure 1.

**Figure 1.**
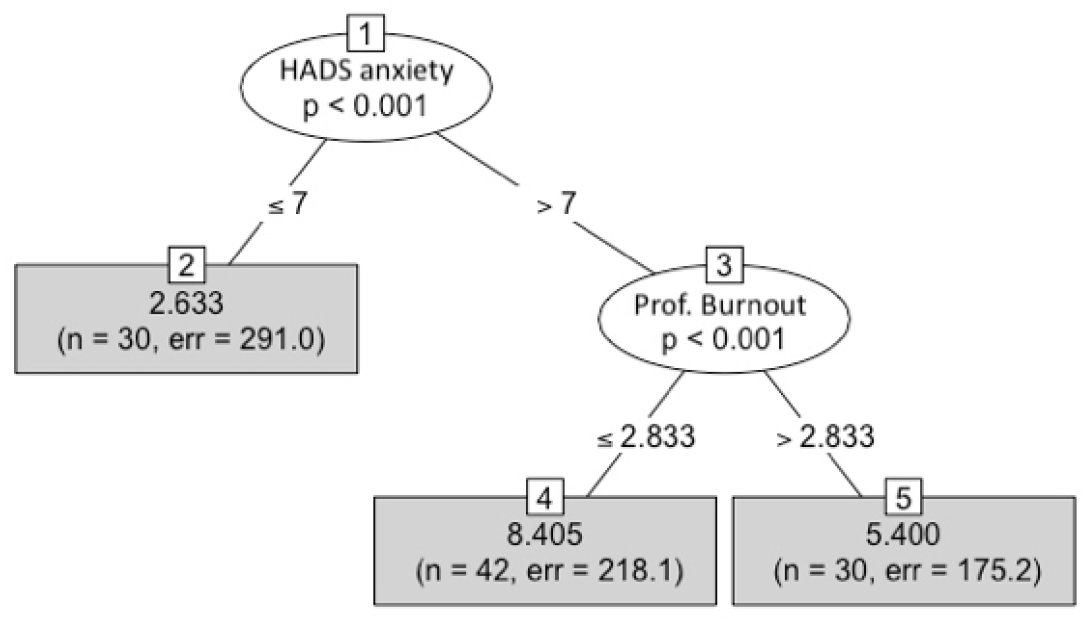
Decision tree predicting depression, including probability of variables and the number of participants belonging to each terminal node (bottom grey panel)

The tree model can be interpreted by starting at the top of the figure, with the first predictor for depression being HADS anxiety. Each branch can then be followed down to the next node until the final node is reached, which shows the mean depression score for the branch. The most promising combination of variables can be seen in panel 4: the combination of anxiety (scores of > 7) and professor burnout (scores of ≤ 2.8) predicts a depression score of 8.4, a result that approaches significance (cut-off: *≥* 9). This outcome confirms the findings from the hierarchical regression model: low levels of professor burnout predicted higher levels of depression.

In summary, depression prevalence was significantly different between groups. At first glance, this result suggests that professional identity could play a role in predicting depression. However, neither subsequent model found this variable to be a significant predictor. Rather, specific variables such as anxiety and burnout with academic staff were reliable depression predictors. Despite high scores in pain catastrophizing in non-musicians and reporting long durations of perceived pain in both musicians’ groups this factor was insignificant in the model.

## Discussion

The aims of this study were to add to our knowledge of how we may better predict depression in students by increasing understanding of how known depression predictors relate to depression experiences in student populations where the main study subject had been evaluated as a variable that might lead to or increase depression. In this study we examined six depression predictors (anxiety, depersonalization, coping strategies, professional identification, pain catastrophizing and burnout).

Following group testing and regression modelling of the data, only two of these factors were found to be significant in predicting depression in our student population: general anxiety and burnout with teaching staff. It was surprising to find that the non-musicians’ group reported significantly more anxiety compared to the musicians’ groups. While this is in line with the literature for medical students (45,46), it goes against expectations from the literature on musicians (47,48). There could be several reasons why both groups of musicians in our study reported lower anxiety levels compared to non-musicians. Firstly, there is a possibility of habituation. Musicians could have grown accustomed to stressful situations and developed a better coping routine. Anxiety may only peak in high-stress situations, such as right before a performance, and then descend to a lower anxiety base level right after, a working hypothesis that could be tested in future. Secondly, it is possible that due to the years of playing successfully in concerts and auditions, musicians evaluate anxiety-inducing situations differently than others, and dispose of greater self-efficacy compared to non-musicians, another hypothesis that is suited to further testing. Thirdly, we should also not discount the possibility that making music could have a long-term therapeutic effect on active musicians. Making music might offer an outlet and the possibility to channel anxiety. The fact that, despite lower anxiety prevalence, college musicians showed higher depression scores, does not decrease the importance of anxiety as a depression predictor. Instead, this result shows that depression is multifactorial and requires a combination of several predictors alongside an assessment of anxiety. The high depression scores for music college musicians might have been influenced by stress from one or several other predictor variables that were not accounted for (49,50).

Burnout only became a depression predictor when combined with anxiety. Here, only a low burnout level with professors (≤ 2.8) predicted depression. This contrasts the data found in reference to depression predictors anxiety and pain catastrophizing, where a higher level predicted higher depression (51,52). How should this disparity be explained? In this context, it is vital to note that the World Health Organization has revised its approach to burnout as a condition in its own right from 2022 onwards (53). If both conditions coincide but do not fully overlap it could explain why a lower level of burnout predicted a higher level of depression. Studies with college athletes found specifically sports-related stressors, such as perceived conflict with professor/ trainers to be the significant predictor for burnout (55,56).This might be one explanation why professor burnout had a greater impact as depression predictor than work or student burnout. It is a further indication for the importance of the (teaching) environment in both conditions.

This was the first study to explore whether professional musical identity could be considered as a valid predictor for depression. Contrary to expectations, our results showed that it was not a significant predictor for depression here. Given both musicians’ groups’ identity scores, and in view of the parallel findings in college athletes, it is reasonable to assume as a future hypothesis that high levels of subscales self- and social identification have a positive impact on depression (i.e. no predictive power), while exclusivity and negative affectivity have a negative impact in depression (i.e. higher score in one predicts higher score in the other) (57). Based on this result, we can conclude that individuals belonging to a certain profession should not automatically be associated with a certain level of depression. Thus, depression research should focus on understanding generalised stressors that may impact them in the same way as people in many different professions. One such consideration, derived from our results in professor burnout, is the type of study environment. The higher depression scores for music college musicians compared to university musicians in our study might be explained by a different career focus or institution-specific environmental stressors. Extrapolating from studies with college athletes, where this type of research has been done, their identity level was based on how much they were immersed in sports, but also fed by the environment (57). As similar studies are missing for music colleges, we can only say that stressors for university musicians seem to be fewer compared to those encountered by college musicians and university non-musicians.

Taken together these results have two implications on future research in this area:

1. the variability of depression between student groups, the differentiability of identity with the profession and the high depression scores in non-music/science students suggest that further interdisciplinary research is crucial. Contrasting student groups not only allows for a better differentiation of depression and depression predicating variables, but also enables to weigh depression factors and isolate specific factors.
2. The variability between higher education institutions demonstrated their vital role in students’ experience of depression. Further research is needed to understand the characteristics within various institutions that impact students in terms of depression.

With regards to limitations, it is important to note that despite providing more than the number of participants determined by the a priori power for meaningful results, the overall sample size is modest. Secondly, while this is in keeping with the literature (58), some students chose not to disclose some information on their mental health. Thirdly, for anonymity reasons, variables such as gender, socio-economic background were excluded. The inclusion might have provided some additional information, but on balance anonymity was deemed more important. Fourthly, this study was designed based on the empirical findings in the area of musicians’ pain and depression. We did not anticipate that our findings would contrast most of the current literature. We would suggest that future studies take this into account as they explore this area in more detail.

Still, our findings provide several implications for the field of depression research in students. Depression should thus be understood based on its multifactorial model, and the corresponding predictors such as anxiety, coping strategies and pain processing, rather than being considered innate to a profession. The higher levels of anxiety and pain catastrophizing for science students showed that these were more problematic in this student group rather than university musicians or music college musicians. It also demonstrated that pain processing needs to be assessed since pain perception alone does not allow for a meaningful conclusion regarding depression. Drawing from previous research with athletes (56), this study suggests that depression and burnout only explain each other to a certain degree. Therefore, they should not be used interchangeably. It is more likely that burnout acts as a mediator/moderator rather than as a single or individual predictor for depression. Finally, identifying depression predictors for specific main study subjects could not only change the self-stigma of students (59), but also deliver implications for primary depression prevention strategies in higher education.

This research received no specific grant from any funding agency in the public, commercial or not-for-profit sectors.

### Contribution

MK led the conception and design of the study, data acquisition, analysis, and interpretation. MK and DC drafted the article. DC and RW contributed to the design and conception and to the interpretation of the data, and critically reviewed the article. RT assisted with the data acquisition, data analysis and interpretation, and critically reviewed the article. VJW helped with the conception, design, data acquisition and analysis, as well as the interpretation of the data, and critically reviewed the article. All authors approved the submitted version. The corresponding author attests that all listed authors meet authorship criteria and that no others meeting the criteria have been omitted.

## Data Availability

All data produced in the present study are available upon reasonable request to the authors

